# Seasonal variation and interspecies dynamics among *Plasmodium falciparum* and *ovale* species in Bagamoyo, Tanzania

**DOI:** 10.1101/2025.03.12.25323778

**Authors:** Kelly Carey-Ewend, Aidan Marten, Julia Muller, Editruda Ernest Peter, Melic Odas, Msolo Credo Dominick, Meredith Muller, Srijana Chhetri, Kano Amagai, Isaack Rutha, Fatuma Kisandu, Lusekelo Beka, Oksana Kharabora, Zachary R. Popkin-Hall, Jeffrey Bailey, Jessie K. Edwards, Emily W. Gower, Jonathan J. Juliano, Billy E. Ngasala, Jessica T. Lin

## Abstract

**Background:** Malaria control in sub-Saharan Africa is typically focused on *Plasmodium falciparum* (*Pf*), but non-falciparum species like *P. ovale curtisi* (*Poc*) and *P. ovale wallikeri* (*Pow*) appear to be rising in prevalence, especially in East Africa.

**Methods:** We conducted polymerase chain reaction (PCR)-based screening of 7,173 asymptomatic individuals over 5 years of age in coastal Tanzania from 2018-2022, employing real-time 18S rRNA PCR assays for *P. falciparum* and *P. ovale*, followed by *Poc*/*Pow* detection. *Plasmodium* positivity was compared across seasons and demographic groups, and interactions between species were analyzed via binomial regression.

**Results:** *Pf* infection (prevalence 27.4%) was associated with younger age, male sex, and higher recent cumulative rainfall, whereas these associations were not apparent for *P. ovale* (*Po*, prevalence 11.5%). *Po* infections appeared to peak during months with lower *Pf* prevalence, especially during the long wet season, when *Po* mono-infections predominated and fewer *Pf*-*Po* co-infections were detected than expected by independent assortment. This apparent antagonism was reversed during the short wet season: *Pf*-*Po* co-infections were comparatively enriched despite low overall *Po* prevalence. In contrast, excess mixed *Poc*/*Pow* infections were detected across all seasons, composing 23% of the *Po*-positive isolates in which a specific *Po* species could be detected.

**Conclusions:** The epidemiology of *P. ovale* species in coastal Tanzania suggests they are frequently present when *P. falciparum* recedes, but also co-infect the same hosts during the short wet season. Meanwhile, the individual *Poc* and *Pow* species often co-exist within individuals, perhaps due to co-transmission or concurrent relapse.

## Introduction

Four *Plasmodium* parasites - *P. falciparum (Pf)*, *P. malariae*, and *P. ovale* (*Po*) species *Po curtisi* (*Poc*) and *Po wallikeri* (*Pow*) - cause most malaria across sub-Saharan Africa, each in dynamic interplay as they co-circulate in the same communities. Prior observational studies, challenge infection experiments, and mathematical models have demonstrated variable propensity for multiple *Plasmodium* spp. to co-exist within individual hosts and vectors, and various intersecting mechanisms are thought to influence these patterns [1–11]. Excess detection of multi-species *Plasmodium* infections has been attributed to carriage within the same vectors, immune suppression, activation of relapses, and modulation of within-host niches (for example, anemia caused by *Pf* leading to reticulocytosis promoting *P. vivax*) [3,6,9,12–14]. Known heterogeneity in biting exposure and malaria risk may also lead to frequent co-infections among particular individuals in a given population [15]. Conversely, mutual suppression by within-host competition, immune protection, and differential distributions across vectors likely contribute to distinct detection of different *Plasmodia* across hosts [11,16,17].

Characterizing patterns of *Plasmodium* interaction is especially important given the changing landscape of malaria in sub-Saharan Africa. Transmission of *P. falciparum*, the primary cause of clinical malaria in the subcontinent, has declined in many regions and is generally much lower than the historical baseline [18]. Meanwhile, non-falciparum malaria spp., including *P. ovale* spp. infections, appear to be on the rise in some of the same regions where falciparum malaria has declined [6,18–21]. This recalls the persistence of *P. vivax* in regions where falciparum malaria has largely been eliminated, suggesting non-falciparum spp. may fill an ecological niche left by *P. falciparum* [22,23]. Since *P. ovale* spp infections are often low-density and subclinical, molecular screening is best suited to investigating these patterns.

We previously characterized asymptomatic *P. falciparum* carriage in Bagamoyo, Tanzania, showing considerable heterogeneity of transmission in an area where *P. falciparum* transmission has fallen but submicroscopic parasitemia remains prevalent [24]. We also reported substantial prevalence of *P. ovale* at our study site over the first three transmission seasons [25]. Here we present full data from more than three years of prospective cross-sectional PCR screening of roughly 7,000 individuals for *P. falciparum* and *P. ovale*, including *P. ovale* spp. identification, to investigate the epidemiology of co-circulating *Pf, Poc,* and *Pow* in East Africa.

## Methods

### Project TranSMIT

Participants aged 6 and older in the Bagamoyo district of Tanzania were recruited from schools and health clinics for cross-sectional malaria screening as part of the Transmission of Submicroscopic Malaria in Tanzania (TranSMIT) study [25,26]. Malaria transmission in this region occurs year-round, peaking during and after the long *masika* and short *vuli* wet seasons. Overall *P. falciparum* transmission has declined substantially compared to 20 years ago [27]. Recruited individuals reported no fever or antimalarial use in the last seven days. Following informed consent, dried blood spots (DBS) were collected alongside demographic survey data. This study was approved by institutional review boards at the University of North Carolina (ID 276606), Tanzania National Institute for Medical Research (NIMR/HQ/R.8a/Vol.IX/3150), Ifakara Health Institute (IHI/IRB/33–2018), and Muhimbili University of Health and Allied Sciences (MUHAS/DA.282/298/01/C).

### Molecular Screening

DNA was extracted from DBS using a chelex protocol prior to molecular amplification [28]. qPCR assays designed to amplify the *pf18S* rRNA gene in *P. falciparum* (*Pf*) [26] and the *po18S* rRNA gene in *P. ovale* (*Po*) spp. [28] were performed on-site in Bagamoyo using Sahara Hot Start PCR Master Mix (Chai Biotechnologies, Santa Clara, CA, USA) on portable Chai open qPCR machines, except for *Po* assays from samples collected in 2018-9 which were shipped to UNC for molecular detection. The *Pf* assay included a dilution series of positive control mock blood spots with known parasitemia (*Pf*) to enable estimation of parasite density, while the *Po* assay used *po18S* plasmid controls (MRA-180; BEI Resources). Positivity for *Pf* and unspecified *Po* spp. are defined in subsequent analyses as any amplification detected in either assay within 45 cycles. To eliminate false-positive *Po* detection due to cross reactivity in the qPCR assays, the *Po* detection threshold was adjusted down to ≤ Ct 42 in samples with measured *Pf* density over 100 parasites/μL [29].

Remaining DBS were shipped to the University of North Carolina at Chapel Hill, where extracted DNA was employed in qPCRs designed to amplify species-specific portions of the *po18S* rRNA gene between *P. ovale curtisi* (*Poc*) and *P. ovale wallikeri* (*Pow*) [30]. Determination of *Poc* and *Pow* positivity was based on the algorithm described in *Potlapalli* et al. [30] to avoid cross-reactive false-positive mixed infections.

### Statistical Analyses

Study data was stored and managed using REDCap (Research Electronic Data Capture) tools hosted at the University of North Carolina at Chapel Hill [31], analysis was performed using SAS software v9.04.01M7P080520 (SAS Institute Inc., Cary, NC, USA) with an α of 0.05, and data visualizations were produced in SAS or R v4.3.2 [32].

We visually investigated the relationship between age and *Pf*- and *Po*-positivity using locally-estimated scatterplot smoothing (LOESS) plots. We then used log-binomial regressions to estimate *Pf*/*Po* positivity by various functional forms of age and rainfall and compared them by Aikake Information Criterion (AIC). AIC and visual fit to LOESS curves led age to be coded categorically with breaks at 11yo and 15yo for downstream analyses. We similarly investigated the functional form of rainfall (mm/day, drawn from the CHIRPS database [33]), including lags of 6, 9, and 12 weeks (w) from date of rainfall to date of screening. This led to selection of restricted quadratic splines of 6w-lagged difference in average daily rainfall over the last month compared to the previous 3mo. The long wet season and short wet season are defined as March-May and October-December, respectively, with a 6w lag to screening date [34].

We modeled positivity for *Pf*, any *Po* species, *Poc*, and *Pow* as binary outcome variables for bivariate analyses examining their distributions by sex, age categories, season, and region. Region is defined as location of self-reported village of residence in the South or North of the district (**Supplementary Figure 1**). We reported proportions alongside Wald asymptotic confidence intervals and compared to demographic variables with Pearson’s *X*^2^. We performed an indirect standardization of wet season *Po* and *Pf* prevalence to the sex and age (in 5-year bins) population structure of Bagamoyo per the 2022 census [35]. We employed the Wilcoxon rank sum test to compare *po18S and pf18S* Ct values and the Kolmogorov-Smirnov test to evaluate differences in *Pf*/*Po* parasite density distributions between mixed and mono-infections. We calculated association between monthly *Po* and *Pf* prevalences using the Pearson correlation coefficient.

**Supplementary Figure 1.**
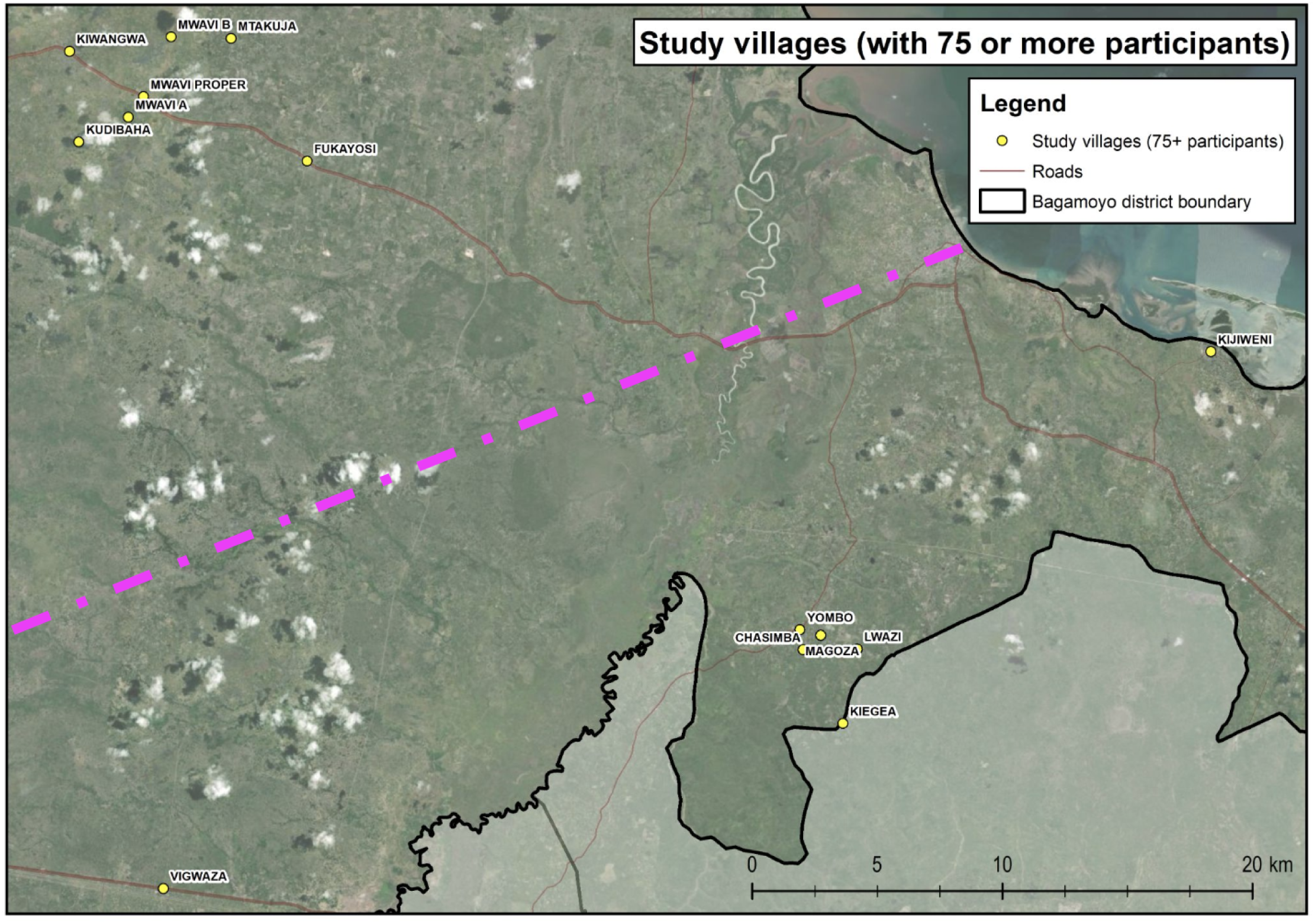
Map of Bagamoyo district and villages with >75 participants screened. Dashed purple line shows the delineation between villages considered in the North and South of the district. Precise GPS coordinates for three villages in the North (Kwa Mkorea, Mwanamvuli, Zemba) and one in the south (Kisumbi) were not available. Reproduced from Rapp et al., *JID* 2024 [24].

To assess adequacy of an interaction-free compartmental model to explain differential *Plasmodium* spp. detection, we fitted the raw data distributions to both a noninteracting distinct pathogens (NiDP) model (which assumes independent transmission of pathogens over the life course) and a multinomial model of independent assortment [36].

We then employed multivariate log-binomial regressions to estimate *Po* prevalence between individuals with and without *Pf* parasitemia. Interaction terms between season and *Pf* status were also included to allow estimated prevalence ratios to vary between seasons. We similarly modeled *Poc* prevalence by *Pow* status to estimate the association between positivity for either *P. ovale* species.

To account for bias in the *Poc*-*Pow* interaction caused by possible assay cross-reactivity and unknown *Po* spp. composition of some samples, we performed two sensitivity analyses using conservative mixed infection determination (requiring similar amplification of both species’ qPCR assays) and randomized assignment of unknown samples to mono-infections of either species (see **Supplemental Materials**).

## Results

### Epidemiology of co-endemic *P. falciparum* and *P. ovale*

From October 2018 to June 2022, 7,173 asymptomatic individuals aged 6yrs and older (68% female, median age 18, IQR 11-30) underwent qPCR screening for *P. falciparum* (*Pf*) and *P. ovale* (*Po*) infection (**Figure 1A, Supplementary Table 1**). Screening was non-contiguous with no sampling during 15 of 45 months, including during the COVID-19 pandemic in 2020. Over the sampled period, 1,968 (27.4%, 95% CI: 26.4-28.5%) individuals were positive for *P. falciparum* and 827 (11.5%, 95% CI: 10.8-12.3%) individuals were positive for *P. ovale* spp. After performing an indirect standardization to the sex and age distribution of the Bagamoyo population, the estimated wet season prevalence of *P. falciparum and P. ovale spp.* among the population aged >5yrs was 30.6%, and 11.7%, respectively. Most infections were low-density: an estimated 71% of *Pf* and 83% of *Po* infections had <100 parasites/μL. Compared to when only one or the other was detected, *Pf*-*Po* spp. co-infections had slightly higher parasite densities (**Supplementary Figure 2**).

**Figure 1.**
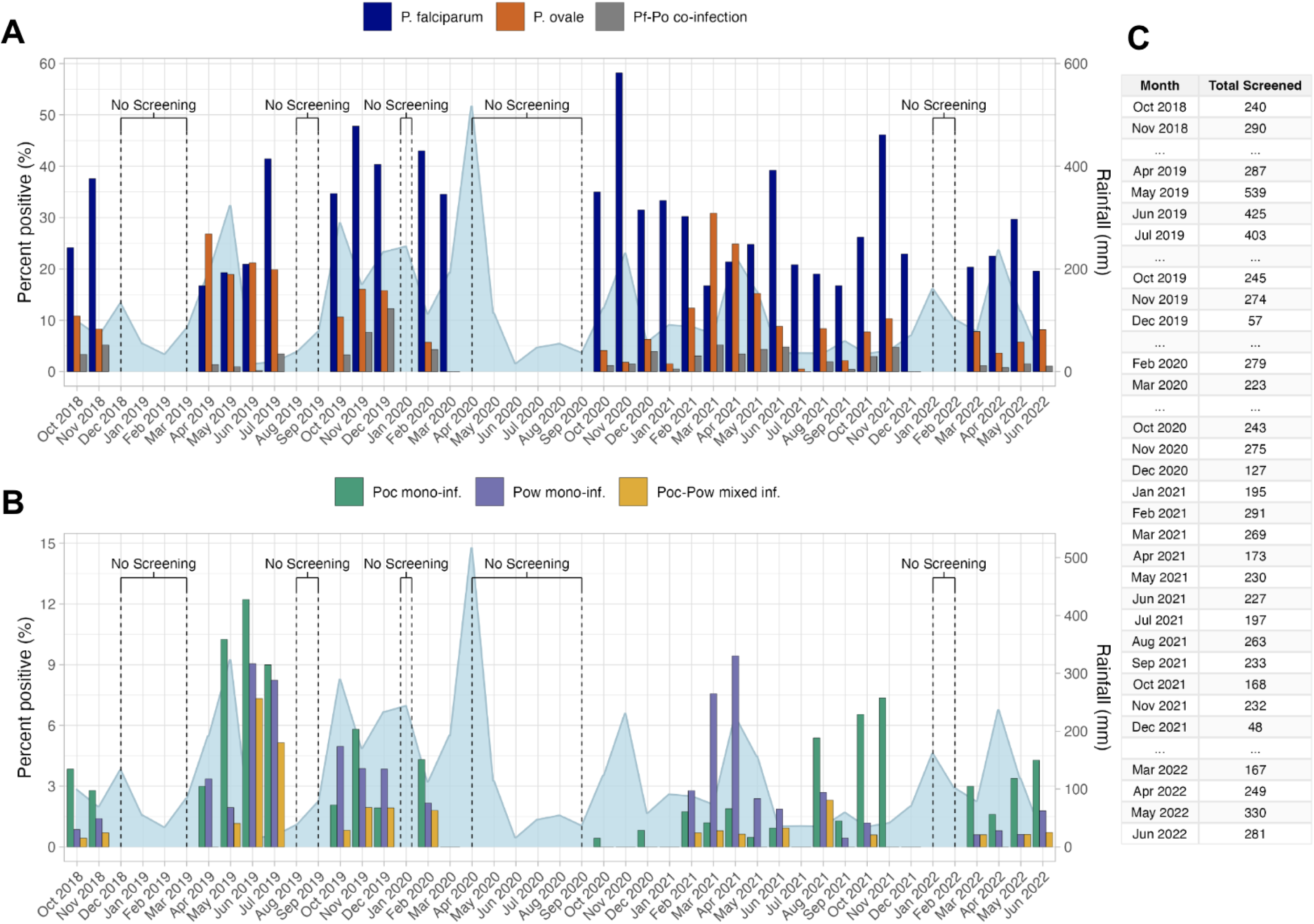
Cumulative monthly rainfall (mm, right) and prevalences (left) of *P falciparum, P ova/e,* and *P ovale-P falciparum* co-infection positivity by month of screening in Bagamoyo, Tanzania, 2018-2022 (A), *P ova/e eurtisi (Poe)* and *P ova/e wa/Ukeri* (Pow) positivity (B), and number of individuals screened per study month (C). Detectable *Poe* mono-infection

**Supplementary Table 1.** Distribution of demographic and molecular screening variables among 7,173 participants screened from 2018-2022 in the Bagamoyo district of Tanzania. Region refers to localization of participant’s self-reported village of residence to the North or South of the Bagamoyo district. *po18S/pf18S* = *Plasmodium ovalelfalciparum* 18S rRNA subunit gene; qPCR Ct= quantitative polymerase chain reaction cyclic threshold; yrs= years; mm= millimeters; Cl = 95% Wald asymptotic confidence interval; **IQR** = interquartile range.

| Variable | Distribution:<br>n (%; CI) or median (IQR) | Number missing<br>data |
| --- | --- | --- |
| *P. ovale* parasitemia | 827 (11.5%, 10.8-12.3%) positive | 1 (0.1%) |
| *P. falciparum* parasitemia | 1968 (27.4%, 26.4-28.5%) positive | 0 (0%) |
| *po18S* qPCR Ct | 40.8 (38.8-42.1) | 3/827 (0.4%) |
| *pf18S* qPCR Ct | 35.8 (31.8-38.5) | 2/1968 (0.1%) |
| Sex | 4853 (67.7%, 66.6-68.7%) female | 0 (0%) |
| Age (yrs) | 18 (11-30) | 1/7173 (0.01%) |
| Average Rainfall<br>(mm/day in past month) | 3.3 (1.5-6.3) | 0 (0%) |
| Region | 3509 (58.6%, 57.3-59.8) North | 1184/7173 (17%) |

**Supplementary Figure 2.**
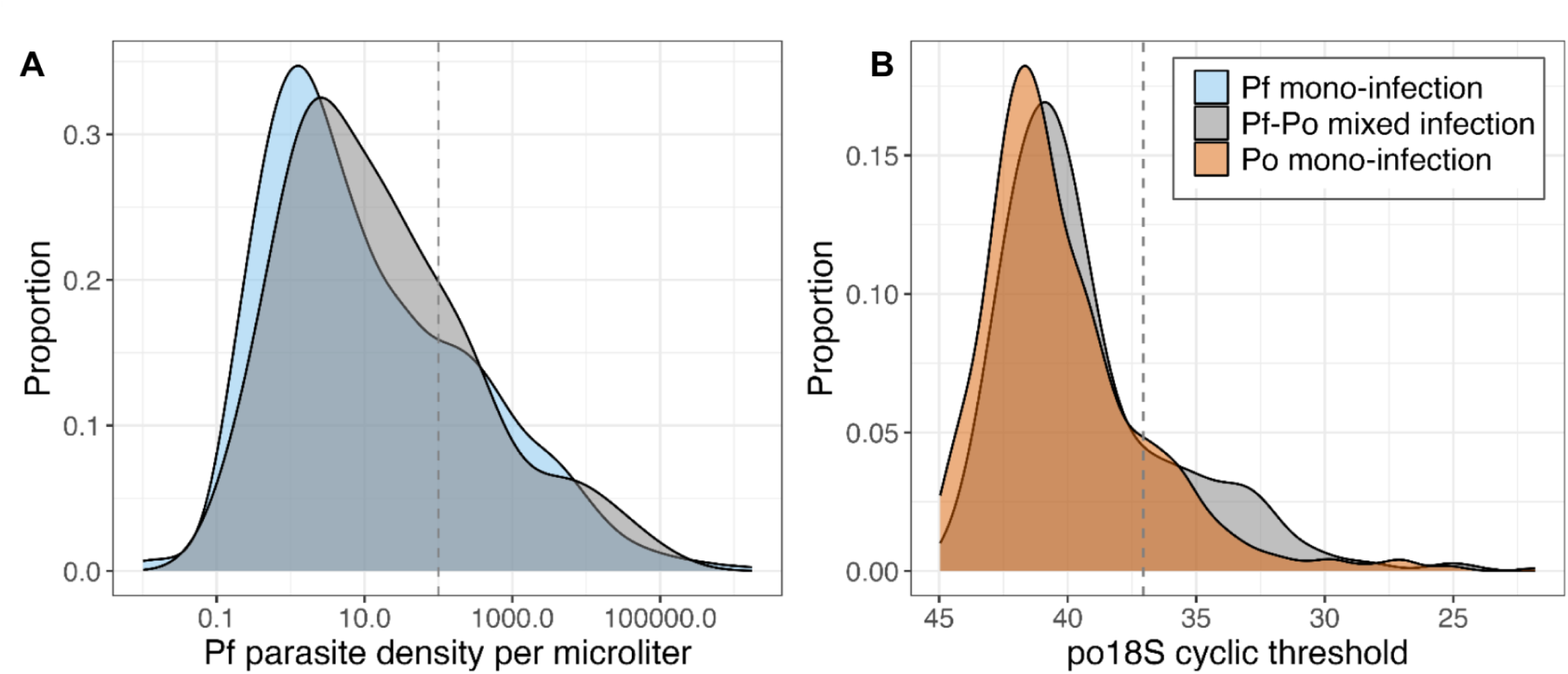
Histograms of parasite density of *P. fa/ciparum* (Pf}-positive samples (A) and *P. ovate* spp. (Po)-positive samples (B) among 1,812 *Pf* mono-infections, 671 *Po* mono-infections, and 156 *Pf-Po* mixed infections. *Pf* parasite density is calculated based on a standard dilution series amplified in the same *pf1BS* PCR (polymerase chain reaction) assay as each sample, whereas *po18S* cyclic threshold (Ct) is presented as a proxy for *Po* parasite density because a *Po* standard series was not tested in each assay (low Ct corresponds to high density). Histogram bars are colored by presence or absence of the other *Plasmodium* parasite. Dashed vertical line depicts the submicroscopic threshold (100 parasites/µL}; for B, this value was imputed from a *po18S* standard dilution series run separately. Parasite density distributions were different between mixed and mono-infections for both *Pf* and *Po* (p-values = 0.030 and 0.013, respectively).

*Pf* and *Po* prevalence varied seasonally and across study years, often in an oscillating manner (**Figure 1**). For example, *Pf* was most common in the short wet season (39% prevalence) and least in the long wet season (24% prevalence), while *Po* showed the opposite pattern (15% and 5% long and short wet season prevalence, respectively) (**Table 1**). Additionally, *P. ovale* prevalence appeared greater in periods with more modest *Pf* prevalence, and dropped when *Pf* transmission was greater, such that monthly *Pf* and *Po* prevalence were negatively correlated (r = -0.23, p = 0.21) **(Figure 2A**). While *Pf* prevalence increased after periods of more rainfall, we were unable to detect a similar trend for *Po* (**Figure 3A**).

**Table 1.**
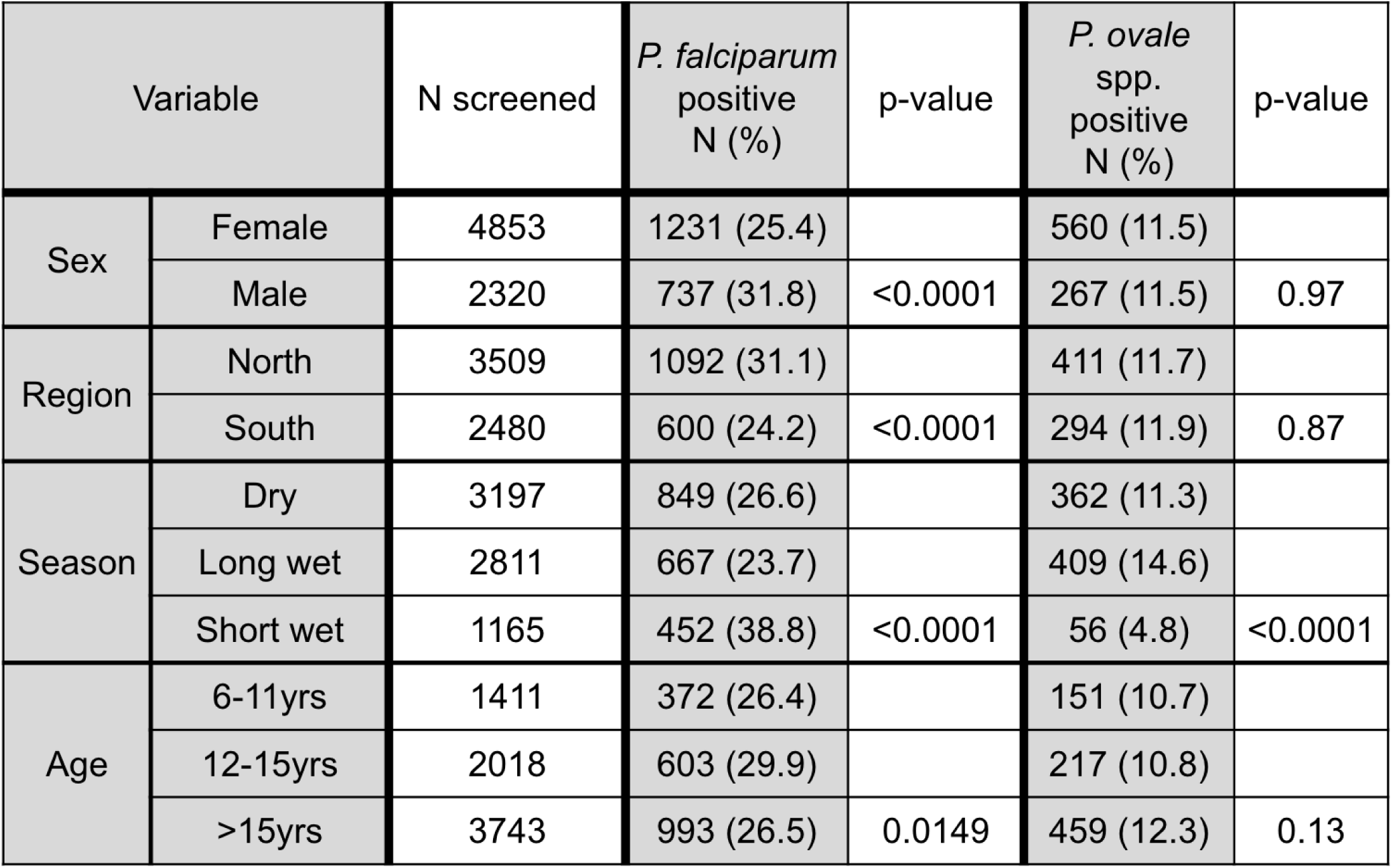
Unadjusted association of demographic characteristics to *P. falciparum (Pf)* and *P ovale (Po)* positivity among 7,173 study participants. Region refers to the relative location of reported village of residence in the Bagamoyo district. Season refers to the long (March-May) and short (October-December) wet seasons incorporating a six week lag. Association of *Pf/Po* positivity with demographic variables is tested using Pearson’s *X^2^*.

| Variable |  | N screened | *P. falciparum*<br>positive<br>N (%) | p-value | *P. ovale*<br>spp.<br>positive<br>N (%) | p-value |
| --- | --- | --- | --- | --- | --- | --- |
| Sex | Female | 4853 | 1231 (25.4) |  | 560 (11.5) |  |
|  | Male | 2320 | 737 (31.8) | <0.0001 | 267 (11.5) | 0.97 |
| Region | North | 3509 | 1092 (31.1) |  | 411 (11.7) |  |
|  | South | 2480 | 600 (24.2) | <0.0001 | 294 (11.9) | 0.87 |
| Season | Dry | 3197 | 849 (26.6) |  | 362 (11.3) |  |
|  | Long wet | 2811 | 667 (23.7) |  | 409 (14.6) |  |
|  | Short wet | 1165 | 452 (38.8) | <0.0001 | 56 (4.8) | <0.0001 |
| Age | 6-11yrs | 1411 | 372 (26.4) |  | 151 (10.7) |  |
|  | 12-15yrs | 2018 | 603 (29.9) |  | 217 (10.8) |  |
|  | >15yrs | 3743 | 993 (26.5) | 0.0149 | 459 (12.3) | 0.13 |

**Figure 2.**
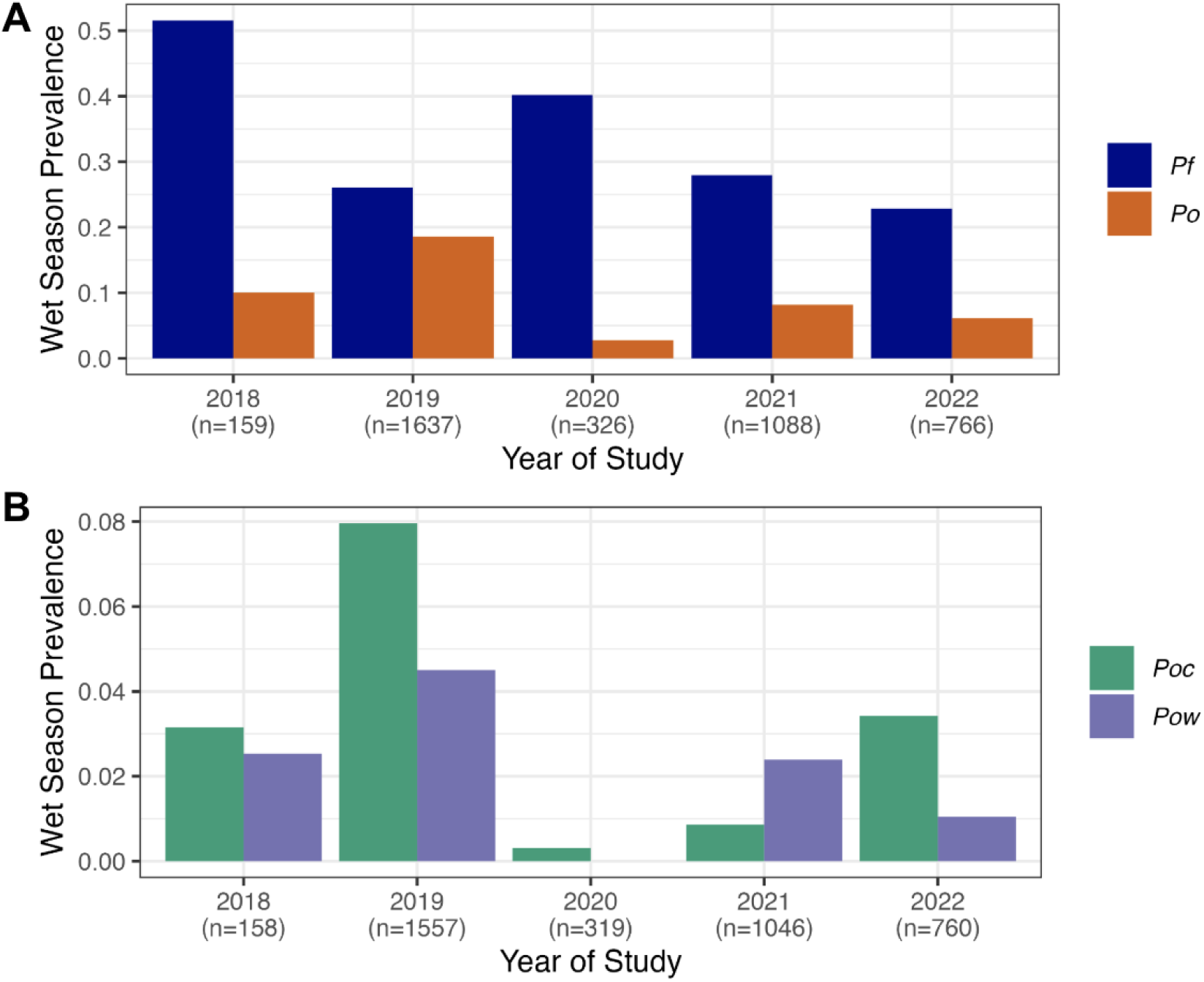
Prevalences of *P. falciparum (Pf)* and *P. ovate (Po)* (A) and detectable *P. ovale curtisi (Poe)* and *wallikeri (Pow)* (B) in the wet season (combined short and long) across study years. Y-axis range is reduced in (B) to show lower prevalence.

**Figure 3.**
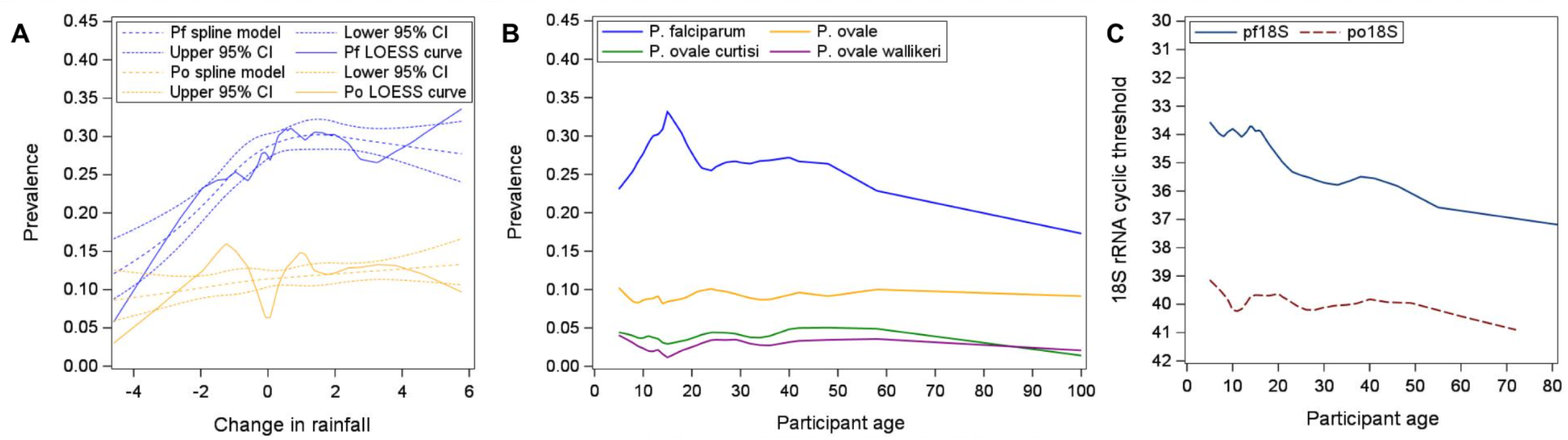
Prevalence of *P. falciparum (Pf)* and *P. ovate* by change in local rainfall (A) and participant age (B) among 7,460 screened individuals, and 18S rRNA polymerase chain reaction (PCR) Ct values by age among 826 and 1968 *Po-* and Pf-positive participants, respectively (C). Association of age is also shown by detectable *P. ovate curtisi* and *wallikeri* prevalence. Change in rainfall is coded as the difference in average rainfall (mm/day) between the preceding 1 month (mo) and the preceding 3mo, starting 6 weeks prior to screening. LOESS smoothed prevalences across the age and rainfall distributions are calculated using the 25% nearest observations; rainfall-prevalence relationships are also modeled using restricted quadratic spline models with breakpoints drawn at quartiles (alongside 95% confidence intervals [Cl]).

Demographic characteristics associated with parasite carriage also differed between species. While *Pf* infections were more common among males and in older school-aged children (12-15yrs), *Po* infections were evenly distributed across sex and all age groups (**Table 1**, **Figure 3B**). These patterns persisted when examining *Po* infections without *Pf* co-infection. *Pf* parasite densities generally decreased with increasing age, while *Po* densities were low across all age groups (**Figure 3C**). Finally, while *Pf* was more prevalent in the northern part of the study area, *Po* spp. did not show any geographic structure (**Table 1**). *Pf* prevalence varied across different villages, ranging from 15-45%, but there was no association with *Po* prevalence (correlation of *Pf* and *Po* village-specific prevalence r = 0.04, p = 0.87) (**Supplementary Figure 3**).

**Supplementary Figure 3.**
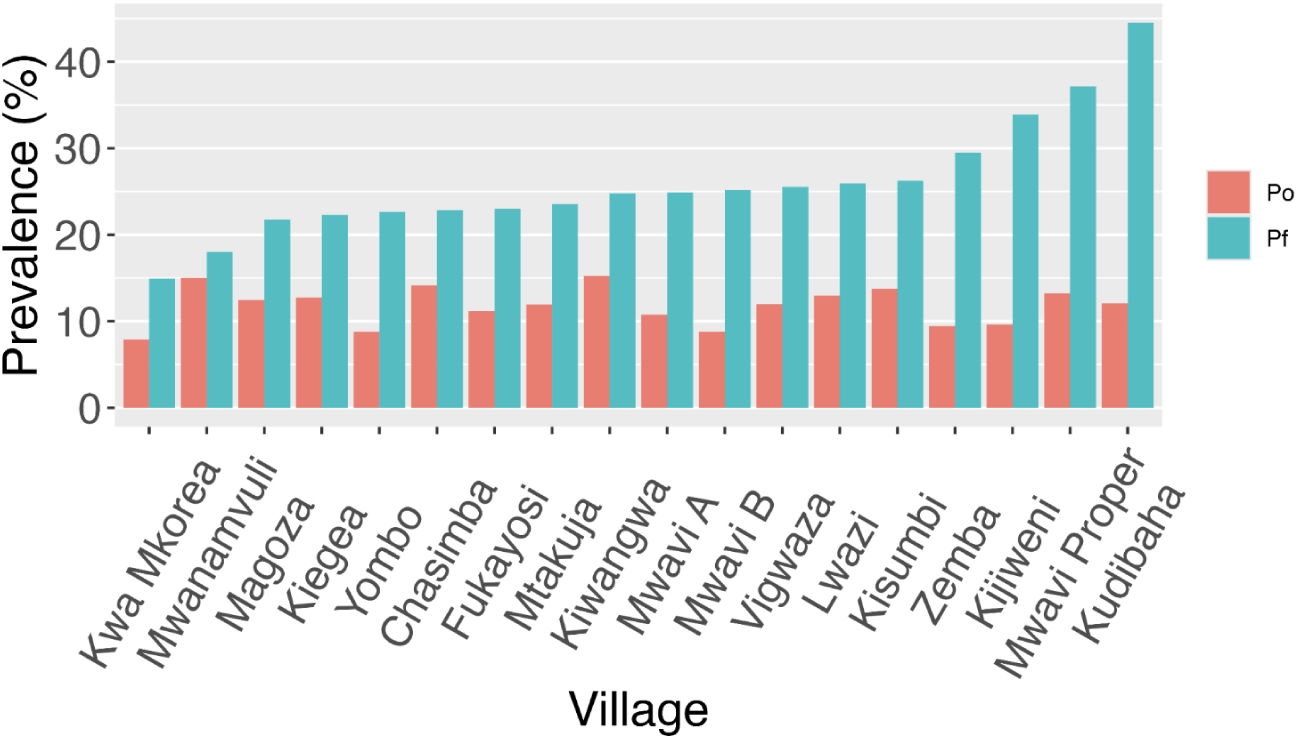
*P. fatciparum (Pf)* and *P. ovate* (Po) prevalences among the 18 villages with >75 participants screened. Villages are ordered by ascending *Pf* prevalence.

### Seasonal dearth of *P. falciparum - P. ovale* spp. co-infection

Reflecting the contrasting epidemiology described above, the number of *Pf*-*Po* co-infections was lower than would be expected by independent assortment of the two *Plasmodia* in the population. Given a *Pf* prevalence of 27.4% and a *Po* prevalence of 11.5%, we would expect to detect 227 *Pf*-*Po* co-infections among 7,173 participants by independent assortment, but we observed 156 co-infections, 69% of the expected value. The data distribution also could not be explained by use of a noninteracting distinct pathogens (NiDP) model [36]. After adjusting for year of study, participant age, rainfall, and season, individuals positive for *P. falciparum* had 0.48 times (95% CI: 0.39-0.60; p < 0.0001) the probability of also being *Po*-positive compared to individuals without *Pf* infection. This antagonism was most pronounced in the long wet season (*Po* PR: 0.24, 95% CI: 0.14-0.40, p < 0.001), present in the dry season (*Po* PR: 0.79, 95% CI: 0.63-0.99, p = 0.04), and reversed in the short wet season (*Po* PR: 1.58, 95% CI: 0.77-3.25, p = 0.21) (**Figure 4A**). The ratio of *Po* mono-infections to *Pf-Po* co-infections in the short wet season were correspondingly low across study years, while long wet season ratios ranged significantly higher (**Figure 4D**).

**Figure 4.**
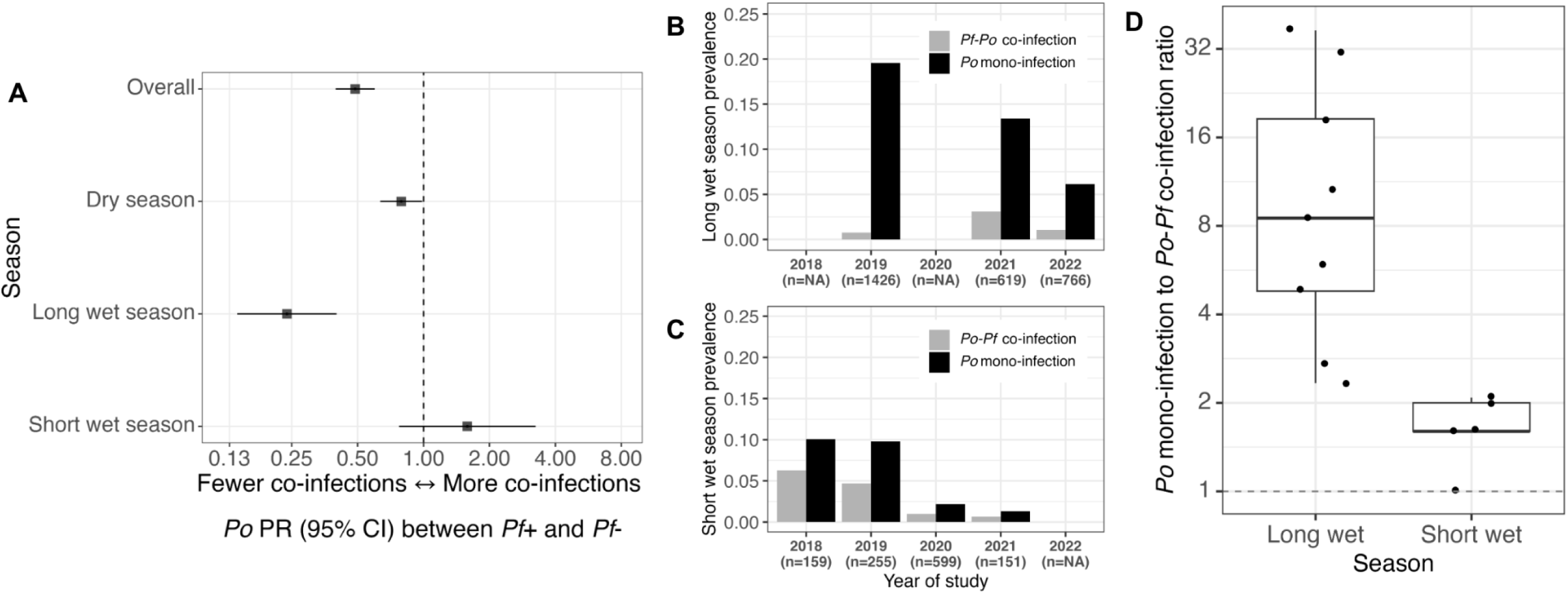
Seasonal variation in *P. falciparum (Pf)* detection among *P. ovale (Po)* carriers. Adjusted *Po* prevalence ratios (PR) by Pf co-infection status (A) and comparison of *Po* mono-infection and *Po-Pf* co-infection (B, C, D) by season. Log-binomial regressions compare *Po* prevalence between PApositive and negative individuals, overall and among different seasons, after adjustment for age, difference in rainfall over preceding 1 mo and 3mo, year of study, and season (in the overall estimate) (A). *Po* mono- and *Po-Pf* co-infection prevalences by year (B, C) and ratios by month (D) are split into participants screened 6 weeks after the long (March-May) and short (October-December) wet seasons. Mono-infection and co-infection refer only to presence of *Po* and *Pf.* Boxes in (D) reflect 1st, 2nd, and 3rd quartiles; whiskers extend to highest and lowest values within 1.5*interquartile range; data plotted on log scale.

### Epidemiology of co-endemic P. ovale curtisi and P. ovale wallikeri

Real-time PCR assays successfully detected *P. ovale curtisi* (*Poc*) or *P. ovale wallikeri* (*Pow*) in 376/631 (59.6%) of Po-positive samples tested (out of 827 total *Po*-positive samples). Among these 376, only *Poc* was detected in 189 (50.2%), only *Pow* was detected in 100 (26.6%), and both *Poc* and *Pow* were detected in 87 (23.1%) (**Supplementary Table 2**).

**Supplementary Table 2.** Final species determination among 827 *P ovale* (Po)-positive and 6,346 Po-negative participants. *Poc* mono-infection (mono-inf.) and *Pow* mono-infection reflect samples with amplification of the primer set for only one species or amplification of both species’ primer sets at disparate levels that could be attributed to cross-reactivity in the assays. *Poc-Pow* mixed infections conversely reflect samples with shared amplification of primer sets for both species at sufficiently similar cyclic thresholds to represent simultaneous parasitemia with both *P. ovale* species (see *Potlapalli et al., PLOS NTD, 2023* [30]). The final reflects proportional species composition among 631 Po-positive samples in which the species-identification assays were performed. 255 Po-positive samples did not show amplification of either primer set, and 196 Po-positive participants lacked sufficient remaining dried blood isolate to run the assays.

| *Po* species status | *Po*-negative | *Poc* mono-inf. | *Pow* mono-inf. | *Poc-Pow* mixed | Unknown species | No assay run |
| --- | --- | --- | --- | --- | --- | --- |
| N | 6346 | 189 | 100 | 87 | 255 | 196 |
| % | 88.5 | 2.6 | 1.4 | 1.2 | 3.6 | 2.7 |
| % of tested *Po*+ | N/A | 30.0 | 15.8 | 13.8 | 40.4 | N/A |

*Poc* and *Pow* parasitemia also varied over the course of the study in relation to each other and to *P. falciparum* transmission (**Figure 1B**). While *Poc* and *Pow* prevalences by study month were significantly correlated (Pearson correlation coefficient = 0.43, p=0.019) (**Figure 2B**), some alternating patterns were also present. For example, *Poc* predominated during the waning months of 2018, the latter half of 2021, and the first half of 2022, while *Pow* was the major *P. ovale* parasite found among those screened in the first half of 2021. During the period of highest *Po* spp. prevalence over the long wet season of 2019 (March-May), both *Poc* and *Pow* were detected, with a relative enrichment of mixed *Poc*-*Pow* co-infections in months following (June-July). Conversely, during the period of highest *Pf* prevalence over the short wet season spanning October 2020-January 2021, *P. ovale* prevalence was 1.5-6.3% and only *Poc* was detected.

Otherwise, the two *P. ovale* spp. showed generally similar patterns of detection by season, age, and sex (**Table 2**). *Poc* and *Pow* were both more likely to be detected in the long wet season compared to the dry season, and least prevalent in the short wet season. *Pf* was most common among adolescents, while *Pow* had the lowest prevalence among adolescents compared to children 6-11yrs and adults (p = 0.02) (**Table 2**, **Figure 3B**).

**Table 2.** Unadjusted association of demographic characteristics to *P. ovale curtisi (Poc)* and *P. o. wallikeri (Pow)* positivity among 6,977 participants, and to *Poc* vs. *Pow* status among 287 *Po* spp. mono-infections (only one *Po* species detected). Association of *Po* spp. positivity (among all participants) and status (among mono-infections) with demographic variables is tested using Pearson’s *X^2^*.

| Variable |  | N screened | *P. ovale curtisi* positive N (%) | p-value | *P. ovale wallikeri* positive N (%) | p-value | *P. ovale curtisi* mono-infections (column % out of 187) | *P. ovale wallikeri* mono-infection (column % out of 100) | p-value |
| --- | --- | --- | --- | --- | --- | --- | --- | --- | --- |
| Sex | Female | 4721 | 185 (3.9) |  | 134 (2.8) |  | 124 (65.6) | 73 (73.0) |  |
|  | Male | 2256 | 91 (4.0) | 0.82 | 53 (2.4) | 0.24 | 65 (34.4) | 27 (27.0) | 0.20 |
| Region | North | 3412 | 135 (4.0) |  | 94 (2.8) |  | 91 (56.9) | 50 (58.8) |  |
|  | South | 2405 | 106 (4.4) | 0.40 | 72 (3.0) | 0.59 | 69 (43.1) | 35 (41.2) | 0.77 |
| Season | Dry | 3137 | 111 (3.5) |  | 80 (2.6) |  | 81 (42.9) | 50 (50.0) |  |
|  | Long wet | 2702 | 152 (5.6) |  | 101 (3.7) |  | 98 (51.9) | 47 (47.0) |  |
|  | Short wet | 1138 | 13 (1.1) | <0.0001 | 6 (0.5) | <0.0001 | 10 (5.3) | 3 (3.0) | 0.41 |
| Age | 6-11yrs | 1378 | 52 (3.8) |  | 40 (2.9) |  | 36 (19.1) | 24 (24.0) |  |
|  | 12-15yrs | 1970 | 71 (3.6) |  | 36 (1.8) |  | 60 (31.8) | 25 (25.0) |  |
|  | >15yrs | 3826 | 53 (4.2) | 0.49 | 111 (3.1) | 0.02 | 93 (49.2) | 51 (51.0) | 0.40 |

### Excess P. ovale curtisi and P. ovale wallikeri mixed infection

Unlike for *Pf* and *Po*, the observed prevalence of *Poc/Pow* mixed infections was higher than expected by independent assortment. *Poc* and *Pow* prevalences were 4.0% (289/6977) and 2.7% (187/6977), respectively, with 87 mixed infections observed in comparison to only 7 expected (**Figure 5**). Again, a NiDP model did not show better fit compared to an independent assortment model. Almost half of all *Pow* carriers had detectable *Poc* despite the relatively low prevalence of both organisms. This synergy persisted after adjusting for *Pf* infection status, age, year of study, and season: *Pow*-infected individuals had 15 times (95% CI: 13-18; p < 0.0001) the probability of also being *Poc-positive* compared to those without *Pow* infection.

**Figure 5.**
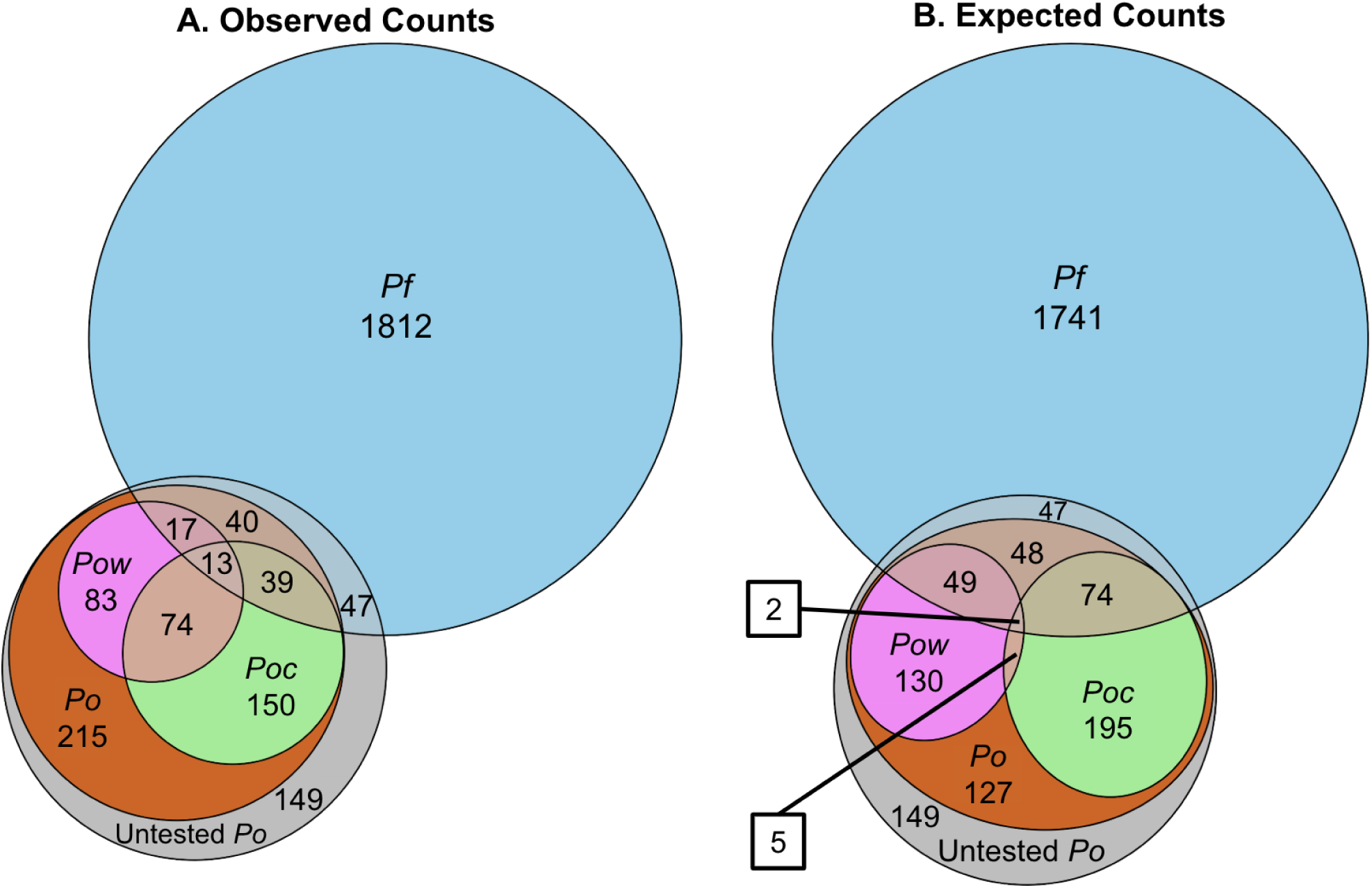
Distribution of observed (A) and expected (B) *P falciparum (Pf), P. ovale (Po),* detectable *P. ovale curtisi (Poc),* and detectable *P ovale wallikeri (Pow)* co-infections among 7173 participants. 196 Po-positive samples were not tested for species composition (Untested *Po).* Expected distributions were calculated assuming independent assortment of marginal positivity probabilities (excluding untested *Po* samples), not including untested samples: P(Pf-positive) = 0.274; P(Po-positive) = 0.090; P(Poc-positive) = 0.040; P(Pow-positive) = 0.027.

To confirm that excess mixed *Poc/Pow* infection was not due to potential cross-reactivity in the 18S rRNA species-identification assays causing false-positivity of the minor species, we reduced the 87 mixed infections to 28 in which the *Poc* and *Pow* assays amplified within 3 cycles of each other; these represent mixed infections in which the minor species exhibits >10% the parasite density of the more prevalent species within the host. Using this conservative dataset, *Pow* carriers still have 3.7 times (95% CI: 3.1-4.5; p < 0.0001) the probability of having detectable *Poc* parasitemia compared to those without *Pow* infection (**Supplemental materials**).

## Discussion

This study provides a comparative portrait of co-endemic *P. falciparum* (*Pf*), *P. ovale curtisi* (*Poc*), and *P. ovale wallikeri* (*Pow*) within an East African community spanning nearly four years and 2,639 persons with asymptomatic *Plasmodium* carriage. We find disparate patterns of infection between *P. falciparum* and both *P. ovale* (*Po*) spp. and seasonal variation in their relative prevalence and degree of within-host overlap, suggesting underlying interspecies dynamics. Specifically, we observe a relative enrichment of *Pf*-*Po* co-infections during the short wet season, as would be expected if transmission relies on the same *Anopheles* vectors. However, this is outweighed by a relative paucity of *Pf*-*Po* co-infections during the rest of the year, with many more *Po* carriers without accompanying *Pf* co-infection during the long wet season. Conversely, among individual *P. ovale* species, we found frequent co-circulation in the same hosts, with marked enrichment of mixed *Poc*-*Pow* infections throughout the year.

We found few prior reports of varying prevalence of different malaria parasites in opposition to each other or with distinct seasonal patterns. *P. malariae* and *ovale* prevalence fell during the wet season when *falciparum* peaked in areas of Burkina Faso, Mozambique, and the Republic of the Congo, though these studies only examined individual years [37–39]. A weekly time series of malaria case data from mountainous areas in Peru showed brief spikes of *P. falciparum* detection that corresponded to troughs in *P. vivax* [40]. However, we could not find longitudinal studies that compared relative species prevalence in the long vs. short wet seasons in East Africa. Our findings may uniquely derive from the serial cross-sectional sampling across years as well as our focus on subclinical malaria.

If the observed seasonal differences in *Pf* vs. *Po* spp. prevalence reflect true infection dynamics in Bagamoyo, they could implicate seasonal differences in abundance of the vectors responsible for their transmission. While the three main malaria vectors in East Africa, *Anopheles gambiae sensu stricto*, *An. funestus s.s.*, and *An. arabiensis* do fluctuate in their relative abundance, with some indication of seasonal alternating predominance [34,41,42] and variable *Pf* carriage [41–44], almost nothing is known about their relative roles as primary or secondary vectors for *P. ovale*. Sporozoite testing from mosquitoes collected in households in Burkina Faso in 2019-2020 revealed a higher *Po* prevalence in *An. funestus s.l.* vs. *Pf* predominance in *An. gambiae s.l*. Our team successfully infected colony-reared *An. gambiae s.s.* via direct skin feeding of *P. ovale*-infected individuals in Bagamoyo, with infectivity that seemed to surpass co-infecting *P. falciparum* in the same hosts [25], but we did not perform the same experiments in *An funestus*. If both *An. gambiae s.l.* (inclusive of *An. arabiensis*) and *An. funestus s.l.* are competent vectors for *Po*, predominance of *An. gambiae s.l.* at the onset of the long wet season and *An. funestus s.l.* near the season’s conclusion, as previously found in Tanzania [34], could lead to reduced *Pf*-*Po* co-infection despite substantial prevalence of both.

During the short wet season, detection of *Pf* and *Po* within the same hosts was higher compared to the other seasons. This finding is more aligned with the frequent *Pf*-*Po* co-infection observed in previous studies in East and Central Africa [4,5,7,8,19,21,45], involving both symptomatic and asymptomatic infections. Aside from abundance of both *Pf* and *Po* vectors in the short wet season, frequent co-infection in this period might derive from activation of *Po* relapses (from latent hypnozoites) by *Pf* super-infection, as is hypothesized to occur for *P. vivax* [14]. Further research into *Plasmodium* inter-species immunity, dynamics of ovale relapse, and vector surveillance for *P. ovale* spp. will be needed to elucidate mechanisms underlying the observed patterns. Worsening climate destabilization and attendant changes in seasonal cycles may add more complexity to deciphering these epidemiological patterns [46].

We successfully identified the specific *Po* species in 376 persons, finding that *P. ovale curtisi* (*Poc*) and *P. ovale wallikeri* (*Pow*) infection patterns largely mirror each other with frequent co-detection in the same hosts. Similar to prior surveys in East Africa [21,47,48], *Poc* slightly predominated, occurring at twice the prevalence among single *Po* spp infections, but *Poc* and *Pow* single-species infections exhibited similar associations with regards to age, sex, and season. Mixed *Poc*/*Pow* infections also appear common in prior small cross-sectional surveys [4,48,49]. This pattern might be attributable to co-transmission in vectors [30], relapse of one *Po* spp. triggered by super-infection of the other *Po* spp., or independent accumulation of both asymptomatic chronic infections over time. Frequent co-detection of *Poc* and *Pow* in the same hosts implies that control efforts, and changes to human and vector populations, may influence both parasite species in tandem.

Our study has multiple limitations. First, while it spans multiple years, screening was discontinuous and not consistent across years and seasons. This is partially addressed in multivariable regressions by adjusting for study year and rainfall. Second, demographic data for all participants were limited to age, sex, and village, and we crucially missed those <6 years of age. Developing immunity in young children may provide unique insights to infection patterns not evident in older age groups [42]. Third, even sensitive PCR assays likely miss low-density infections that circulate at or below their limit of detection [50]; longitudinal sampling probably represents the best strategy for detecting subclinical carriage. Fourth, failure of *Po* species-identification in 40% of positive samples limits our ability to characterize *Po* spp. infections with low parasite density, and assay cross-reactivity from field samples may bias towards detection of excess mixed *Poc*/*Pow* infections, though this was examined with robust sensitivity analyses. Finally, individuals with symptomatic malaria are not included in our survey. It is not known how often individuals with *P. ovale* spp. infection are symptomatic, but P. ovale continues to make up the minority of identified *Plasmodium* species among suspected malaria cases, ranging 0-15% prevalence among 10 Tanzanian regions sampled in 2021 [45]. Interspecies interactions and age-stratification likely manifest differently in symptomatic infections.

This survey of asymptomatic *Plasmodium* infections in East Africa shows seasonal disparities in patterns of infection between *Pf* and *Po* spp. alongside increased detection of *Poc* and *Pow* within the same hosts. These results indicate that *P. ovale* prevalence may be influenced by distinct factors from *P. falciparum* in the same community, including unique vector populations, human immune profiles, and relapse propensity. At the same time, *Poc* and *Pow* inhabit the same hosts without obvious differences in their epidemiology in older children and adults. Therefore, treating falciparum malaria and deploying interventions focused on groups at risk for *P. falciparum* are unlikely to adequately address *P. ovale* burden, while *P. ovale*-specific interventions have the potential to address both *Po* spp. in tandem. Further research that follows *P. ovale* spp. carriers over time, investigates the relationship between asymptomatic and symptomatic infections, uncovers each species’ relapse propensity and periodicity, and explores their clinical impact, will ensure that we pursue appropriate malaria control strategies (including potential treatment of hypnozoites) that do not neglect non-falciparum species.

## Supporting information

de-identified screening dataset

CHIRPS daily rainfall

Poc/Pow-specific parasite density for all tested samples

po18S qPCR assay used for parasite density estimation

## Data Availability

De-identified datasets are provided as supplemental materials. Code for data processing, analysis, and visualization is available at https://github.com/IDEELResearch/TranSMIT_pf-po_epi.

## Supplemental Materials

### Sensitivity analyses employed to investigate excess of mixed infections

We employed a sensitivity analysis to ensure that the inflated probability of *Poc* and *Pow* co-detection within the same samples could not be reasonably explained by cross-reactivity between the 18S rRNA species-specific assays. Such cross-reactivity could present as false-positive low density amplification of the primer set for one species in relatively high density true infections of the other species [30]. While the algorithm we used for *Po* species assignment was designed to have 100% specificity for accurate calling of mixed infections, we performed a sensitivity analysis for potential *Po* spp. misclassification by limiting identification of mixed infections to samples in which the *Poc* and *Pow* qPCR assays amplified within 3 cycles of each other (representing mixed infections in which the minor species’ parasite density represented over 10% the density of the more prevalent species within the host). Samples with more disparate amplification might indicate cross-reactivity and were reclassified as mono-infections of the predominant species in this conservative dataset, before repeating the above statistical analyses to estimate adjusted *Poc*-predominant infection prevalence ratios between individuals with *Pow*-predominant infections vs. individuals without.

An additional sensitivity analysis was performed to ensure that samples with unknown *P. ovale* species composition (*Po*-positive samples in which both species-specific qPCR assays failed) did not explain the observed excess of mixed infections, which would occur if mono-infections were more likely to go unidentified. Over 50% of detected mixed infections were in samples within the top tertile of overall *P. ovale* parasite density (which improves rate of successful species-identification), as compared to only 29% and 27% of *Poc* and *Pow* mono-infections, respectively (**Supplementary Figure 3A**). Maximum expected mixed infections would occur when the population prevalences of *Poc* and *Pow* are equal, so 10 simulations were performed randomizing the unidentified *P. ovale* samples to either *Poc* or *Pow* mono-infections at a ratio of 83:172, respectively, in order to equalize the final prevalences of *Poc* and *Pow*. Adjusted *Poc* prevalence ratios between *Pow*-positive and *Pow*-negative individuals were then calculated in each iteration, yielding 10 estimates.

These two sensitivity analyses were then performed in unison with both the conservative mixed infection identification scheme and randomization of unidentified samples to *Poc* and *Pow* mono-infections, yielding 10 prevalence ratio point estimates.

Using the conservative mixed infection classification approach, *Pow* carriers had 3.7 times (95% CI: 3.1-4.5; p < 0.0001) the probability of detectable *Poc* parasitemia compared to those without *Pow* infection. Among 10 simulations of randomizing the 255 unidentified *P. ovale* samples to *Poc* or *Pow* mono-infections, estimates of the adjusted *Poc* prevalence ratio between *Pow+* and *Pow-* individuals varied from 5.4-5.7 and yielded significant p-values <0.0001 in all iterations. Use of both sensitivity analyses in tandem (conservative mixed infection classification and randomization of missing samples to mono-infections) showed that individuals with *Pow* parasitemia had 1.3-1.4 times the chance of having *Poc* compared to *Pow*-negative participants, with all p-values < 0.05. These indicate that the observed excess of *Poc*-*Pow* mixed infections in this cohort cannot be fully explained by increased assay cross-reactivity nor missing data bias from *P. ovale*-positive samples with unidentified species composition.

**Supplemental Figure 3.**
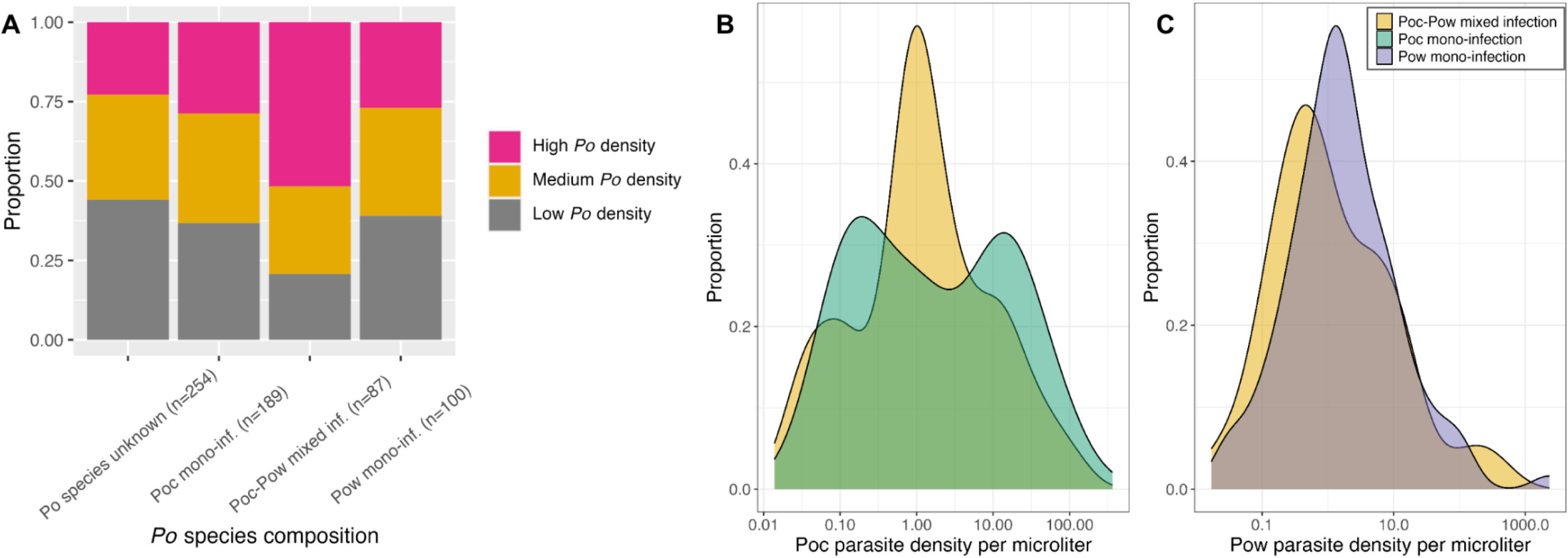
Distribution of overall *P ovate* (Po) parasite density (A), *P ovate curtisi* (Poe) parasite density (B), and *P ovate wallikeri* (Pow) parasite density (C) in *P ovate curtisi* mono-infections (Poe mono-inf.), *P ovate wallikeri* mono-infections (Pow), mixed infections, and infections with unknown *Po* species among 631 Po-positive samples which received both species-identification assays. *Po* density in (A) represents tertiles of all positive *P ovate 18S rRNA (po18S)* gene quantitative polymerase chain reaction cyclic thresholds (Cts), with "high density" referring to the lowest tertile of Ct values. One Po-positive sample was found to be species unknown but lacked *po18S* Ct data and was excluded from this figure. Parasite densities in (B,C) are calculated based on a standard dilution series of plasmid controls amplified in the same assay as each sample. "Mono-infection" refers to an infection with only one *P ovate* species detected, regardless of presence of other *Ptasmodium* species.

## Acknowledgments

We are grateful to the study participants as well as the staff at the schools and health centers in Bagamoyo for their support. We thank the study team at Muhimbili University of Health and Allied Sciences for carrying out the field activities.

## Notes

### Disclaimer

The funders had no role in the study design, data collection, or interpretation.

### Contributions

JTL, BN, and JJJ conceptualized the study. MO, IR, FK, LB collected samples and survey data from participants. TP, DM, MM, SC, AM, JM, KA, and OK conducted molecular laboratory assays and performed data cleaning. KCE and JM performed statistical analyses and data visualizations. KCE and JTL drafted the manuscript. KCE, JTL, JB, EG, JE, JJJ, BN, and ZRPH revised the manuscript. All authors read and approved the final manuscript.

